# Assessing the Seasonality of Lab Tests Among Patients with Alzheimer’s Disease and Related Dementias in OneFlorida Data Trust

**DOI:** 10.1101/2024.03.18.24304494

**Authors:** Wenshan Han, Balu Bhasuran, Victorine Patricia Muse, Søren Brunak, Lifeng Lin, Karim Hanna, Yu Huang, Jiang Bian, Zhe He

## Abstract

About 1 in 9 older adults over 65 has Alzheimer’s disease (AD), many of whom also have multiple other chronic conditions such as hypertension and diabetes, necessitating careful monitoring through laboratory tests. Understanding the patterns of laboratory tests in this population aids our understanding and management of these chronic conditions along with AD. In this study, we used an unimodal cosinor model to assess the seasonality of lab tests using electronic health record (EHR) data from 34,303 AD patients from the OneFlorida+ Clinical Research Consortium. We observed significant seasonal fluctuations—higher in winter in lab tests such as glucose, neutrophils per 100 white blood cells (WBC), and WBC. Notably, certain leukocyte types like eosinophils, lymphocytes, and monocytes are elevated during summer, likely reflecting seasonal respiratory diseases and allergens. Seasonality is more pronounced in older patients and varies by gender. Our findings suggest that recognizing these patterns and adjusting reference intervals for seasonality would allow healthcare providers to enhance diagnostic precision, tailor care, and potentially improve patient outcomes.

## Introduction

In 2023, as many as 6.7 million Americans live with Alzheimer’s disease (AD) [1]. By 2060, this number is projected to double to 14 million, fueled by the aging baby boomers. In 2019, 121,499 deaths from AD were recorded, making AD the 6th leading cause of death among US adults and the 5th leading cause of death among Americans aged ≥ 65 [1]. However, no treatments have been successful in curing or preventing AD. Moreover, many AD patients live with comorbidities such as hypertension (83.6%), respiratory diseases (75.8%), lipoid metabolism disorders (62.2%), joint disorders (53.6%), cataracts (48.8%), and osteoarthrosis and allied disorders (50.5%) [2], that worsen their cognitive decline and overall health.

Managing chronic conditions is extremely important but challenging for patients living with AD. Lab tests are essential in monitoring these conditions, adjusting treatments, and managing symptoms effectively. For example, conditions like hypertension, heart disease, and dyslipidemia are common and can significantly impact Alzheimer’s progression and patient quality of life. Lab tests monitoring lipid profiles, blood pressure, and cardiac markers are crtical for these conditions. Diabetes management is also crucial as it can worsen cognitive decline [3], and vice versa [4]. Lab tests such as HbA1c, fasting glucose levels, and insulin levels are used to manage diabetes. In addition, patients with AD often take many medications at the same time. It is therefore important to use lab tests to monitor the effectiveness and side effects of these medications to ensure that polypharmacy is not adversely affecting the patient’s health. However, even with easy access to lab test results through patient portals, many patients still face challenges with lab results interpretation, because the current practice of lab test reporting often only focuses on presenting a value with a universal reference range, without considering patients’ individualized characteristics.

Seasonality refers to a recurring pattern characterized by regular fluctuations [5]. It is particularly prevalent in healthcare datasets due to seasonal shifts in environmental conditions and human behaviors. For example, in Florida, days in summer typically have longer daylight hours, warmer temperatures, and increased humidity, fostering plant growth and higher pollen levels. High levels of pollen can trigger allergies, potentially influencing associated lab test values such as monocytes. Conversely, during winter, people often spend more time indoors and may alter their dietary habits, potentially influencing lab test values such as glucose.

The reference interval, which indicates the normal range of a specific lab test, is typically derived from tests conducted on healthy individuals following International Federation of Clinical Chemistry (IFCC) guidelines [6,7]. This type of reference interval often fails to consider patients with certain diseases or seasonal changes in lab tests. In clinical practice, the interpretation of lab tests often relies on physicians’ experience and interaction with patients, assisted by the guided reference interval. However, this approach is impractical for large-scale EHR-data-driven research and is infeasible for self-care monitoring tools (such as mobile health applications), which require prompt responses from users [8].

Assessing seasonality in lab tests across manifold cohorts is an essential step in informing personalized healthcare services. In current literature, Muse et al. [9] have conducted a comprehensive seasonality study on a general population of nearly 2 million hospital patients in Denmark and have identified seasonality in a wide array of lab tests. Meanwhile, most seasonality investigations have focused on specific disease areas or functional domains, with a limited number of lab tests being checked [10–12]. For example, Cheung et al. [13] have found significant seasonal components in 13 of 21 pre-selected clinical and laboratory markers among chronic hemodialysis patients. Miyake et al. [14] have studied seven liver function related lab tests and demonstrated notable seasonality in AST and ALT with outpatient data. Regarding patients with AD, current studies have investigated seasonality in cerebrospinal fluid biomarkers [15] and births [16]. To the best of our knowledge, a comprehensive seasonality study in multiple lab tests targeting a large AD patient cohort has yet to be conducted. In this study, we employed an unimodal cosinor model on the AD patient cohort data from the OneFlorida Data Trust to identify seasonal variations in 28 lab tests from 34303 AD patients. The cosinor model is a simple and popular method for analyzing seasonal health data, with a parsimonious number of parameters and straightforward interpretation [9,17]. It typically assumes that seasonal variations in lab test results follow recurring patterns that have one rise-and-fall regularly over the course of a year. Additionally, we created and analyzed subgroups of patients based on demographics including gender and age. We utilized the K-Means [18] clustering algorithm to group lab tests with similar seasonal patterns. Manifold visualization tools were used to summarize and compare the analysis results of each group. Our study demonstrates significant seasonality in several lab tests among AD patients, and different demographics may have different extents of seasonality for the same lab tests.

## Methods

### Data source and cohort selection

The current study used AD patient cohort data from the OneFlorida Data Trust, a centralized patient data repository from the OneFlorida+ Clinical Research Consortium. Its core data contains longitudinal and linked patient records of ∼19 million Floridians from various sources, including Medicaid/Medicare, cancer registry, vital statistics, and EHRs from its clinical partners [19]. The objective of the study is to discover the seasonality variation in hospital laboratory tests based on two different strata, namely age and gender, within the population of AD patients. To select the lab test with possible seasonal variation, we used the list from the study of Muse et al. [9]. All the identified lab test names were then mapped back to LOINC codes using the RELMA (Regenstrief LOINC Mapping Assistant) Database. RELMA (https://loinc.org/downloads/relma/) is a publicly available LOINC database that provides extensive details about lab tests, such as LOINC, test names, and descriptions. A total of 122,669 AD patients were identified with International Classification of Disease Ninth Revision Clinical Modification (ICD-9-CM) code 331.0 and ICD-10 CM codes of G30, G300, G30.0, G301, G308, G30.8, G309, G30.9. We further refined the study cohort as all the AD patients having lab test records within the four-year study period of 2016-2019. A total of 34,303 patients were identified based on these cohort selection criteria. Further, a detailed patient cohort selection overview is shown in Figure 1. Note that to improve the robustness of the statistical model [9], we have selected lab tests if they have at least 100 patients in over 26 weeks of a year (50% of the 52 weeks). We further generated subgroups by gender (male and female) and age group (50-79, >=80), respectively. Finally, the cohort of AD patients and two subgroups based on gender and age respectively were used for fitting the statistical models for seasonality variation.

**Figure 1.**
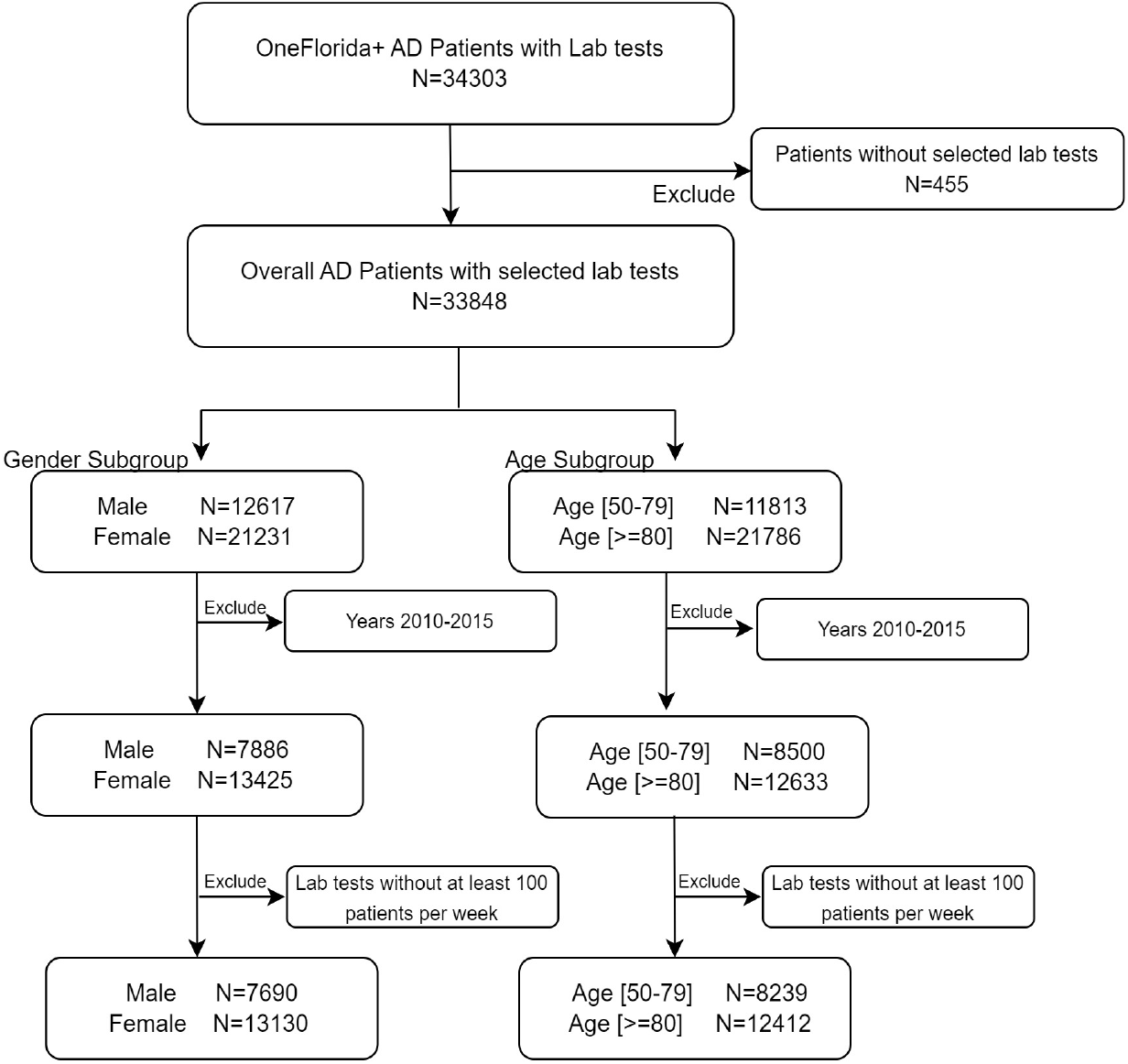
Patient cohort selection process.

### Seasonality analysis

Time-series data consists of a sequence of observations recorded over time, often characterized by two deterministic components, trend and seasonality, and an indeterministic noise component due to sampling variations. The trend reflects long-term developments, while seasonality represents regular fluctuations occurring within a year [20]. In this study, we standardized lab test results by their annual median to eliminate potential trend effects, and then utilized a cosinor model to examine the remaining seasonality component of recurring fluctuations regularly over the course of a year [5]. The cosinor model assumes that lab test fluctuations conform to smooth sinusoidal waves over time, utilizing a cosine function to capture the periodic rise and fall of seasonal variations. This function involves only a few parameters, providing clear control over the amplitude, offset, and cycle of a sinusoidal curve. This simplicity facilitates straightforward interpretation of the results, while still accommodating variations in cycle lengths and shapes with flexibility. The parsimonious parameterization of the model ensures computational efficiency and ease of implementation using standard statistical software packages.

### Cosinor model

Let *y*_*ij*_ be the median of a lab test measured on Week *x*_*ij*_ of the *i*th year, where *x*_*ij*_ *∈*{1, 2, …, 52}. In our study, four years of data were employed. Thus, *i*=1, 2, …, 4 and *j*=1, 2, …, *J*, where *J ∈*{27, …, 52} due to the 50% missingness exclusion criteria. The cosinor model is specified as follows:

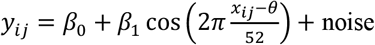

where *β*_0_ denotes the height parameter, indicating the baseline value of the given lab test. Typically, it will be close to zero if observations are normalized to the median before model fitting. The amplitude parameter *β*_1_ quantifies the magnitude of seasonality within the data. It is half of the difference from the highest to lowest point of the sinusoidal wave. This parameter indicates the maximum extent to which a lab test value can fluctuate around the baseline, encompassing both the increases and decreases in values symmetrically. The offset parameter *θ* indicates the week to shift the values’ seasonality. When the amplitude parameter is restricted to the positive region, the offset represents the week in which the variation reaches the yearly peak within a range between 0 and 52. By imposing these bounds, the model is identifiable and has clear and reliable interpretations.

Additionally, one can adjust the cycle of the curve by multiplying a cycle parameter within the cosine term. Since the cycle parameter estimates typically approximate one in our dataset, we chose not to include this parameter to maintain model simplicity.

### Analysis procedure

We conducted analyses on both the full cohort data and subgroups stratified by gender and age, respectively. Lab tests with binary or categorical values were not suitable for the cosinor model and were thus excluded from further analysis. To eliminate the trend over time and effectively assess relative seasonality across years, each observation was normalized to its respective year’s median. This involved subtracting and dividing each observation (*y*_*ij*_) by the median of the *i*th year. The division ensures comparability of amplitude fits across lab tests with varying ranges.

For model fitting, we utilized the R package “nlstools” (version 2.0-1) with the “port” algorithm, which allows for the definition of parameter boundaries [21]. Additionally, each observation was weighted by the number of patients involved in the corresponding week. This weighting approach ensures that observations derived from more patients carry greater influence in the analysis, reducing bias from abnormal records during special events such as holidays [9]. The false discovery rate (FDR) was controlled using the Benjamini-Hochberg method [22]. The K-Means clustering algorithm was used to group lab tests showing similar seasonal patterns within each dataset. This clustering was based on scaled amplitude and offset fits. To account for the cyclic nature of time, offset fits were scaled by measuring their proximity to the 26^th^ week, the summer of a year in Florida.

## Results

### Basic characteristics

The OneFlorida AD patient cohort with the number of lab test records is described in Table 1. The study used 34,303 patients from the OneFlorida Alzheimer’s patient cohort; after applying various conditions, we carefully selected 20,820 (60.69%) patients with AD as the overall cohort to model the seasonality variation. We then applied different strata-based conditions to generate sub-populations as described in Table 2, namely, gender as male (7,690; 22.72%) and female (13,130; 38.78%), and age as 50-79 (8,239; 24.34%) and >= 80 (12,412; 36.67%), respectively. A total of 505 lab tests were selected and mapped with LOINC codes using RELMA to test for seasonality variation. All the lab test LOINC codes were then used to filter the lab tests from the OneFlorida Alzheimer patient cohort.

**Table 1.** Characteristics of the AD patient cohort.

| Characteristics | Number of Patients | Lab Test Records |
| --- | --- | --- |
| OneFlorida Alzheimer | 34,303 | 15,278,707 |
| Selected Labs (505) | 33,848 | 8,783,093 |
| Male | 12,617 (37.28%) | 3,509,285 |
| Female | 21,231 (62.72%) | 5,273,808 |

**Table 2.** Characteristics of two subgroups by gender and age group.

| Subgroup | Characteristics | Number of Patients |
| --- | --- | --- |
| Overall cohort | - | 20,820 (60.69%) |
| Subgroup by gender | Male | 7,690 (22.72%) |
|  | Female | 13,130 (38.78%) |
| Subgroup by age group | Age [50-79] | 8,239 (24.34%) |
| | Age [$\geq 80$] | 12,412 (36.67%) |

Table 3 shows the number of lab tests having at least 100 patients per week for different strata. Among these lab tests, only those lab tests present in all four years of the study period 2016-2019 were used for modeling. This approach gives us the ability to investigate the variation in lab tests across all four years spanning 52 weeks.

**Table 3.** Number of lab tests having at least 100 patients per week for different strata for the study period.

|  | Characteristics | 2016 | 2017 | 2018 | 2019 | Number of Common Lab Tests | Number of Lab Tests for Model Fitting |
| --- | --- | --- | --- | --- | --- | --- | --- |
| Overall | -- | 32 | 32 | 28 | 28 | 28 | 17 |
| Subgroup by Gender | Male | 24 | 23 | 21 | 14 | 13 | 7 |
|  | Female | 26 | 27 | 27 | 25 | 24 | 14 |
| Subgroup by Age Group | Age [$\geq 80$] | 24 | 24 | 24 | 23 | 23 | 14 |
|  | Age [50-79] | 25 | 26 | 25 | 22 | 22 | 14 |

### Seasonality variation

After filtering out lab tests with binary or categorical values, the dataset for the full cohort comprises 17 lab tests for model fitting. For subgroups, the male group includes seven lab tests, the female group includes 15 lab tests, and both age groups include 14 lab tests each (Refer to Table 3 for details). Table 4 presents the fitted results of available lab tests, respectively. After the normalization of observations, the fitted amplitude value 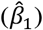 indicates the seasonal fluctuation relative to the baseline value of the lab test. The fitted offset 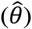 signifies the week of the year when the lab test reaches its peak value. For example, in the case of AD patients older than 80 years old, Eosinophils level is expected to increase by 5.1% around the 29^th^ week, typically during summer, compared to their normal level.

**Table 4.**
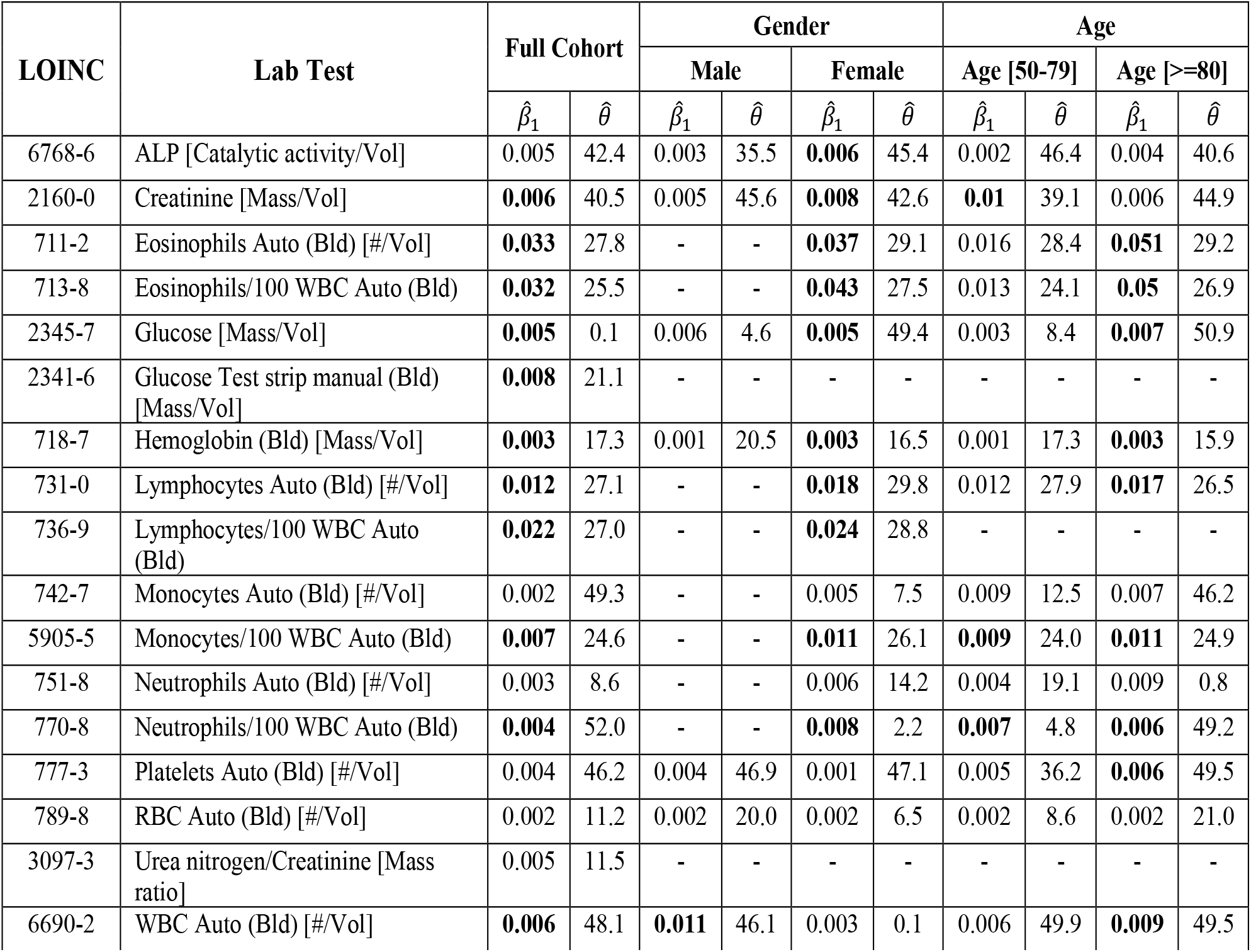
Fitted relative seasonality 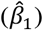 and peaking week 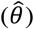 of lab tests for each dataset. Significant relative seasonality fits are in bold. (p<0.05; FDR corrected)

In Table 4, several lab tests have shown significant relative seasonality, indicated in bold. Certain lab tests appeared more pronounced seasonal effects within specific demographic groups. During summer, Eosinophils and Eosinophils/100 WBC increased by over 3% in the overall group, with particularly high fluctuations around 5% in AD patients over 80. Meanwhile, Lymphocytes, Lymphocytes/100 WBC, and Monocytes/100 WBC increased by over 1% across demographic groups, with Lymphocytes/100 WBC showing an increase of over 2%. During winter, seasonal effects were more notable on WBC for males, with an increase of 1.1%. Although these changes may seem small, a previous study found that even slight seasonal variations (1.8% in men and 2.5% in women) in serum total cholesterol concentration resulted in an increase of 22% for the estimated prevalence of hypercholesterolemia (>6.2 mmol/L) [23].

#### (a) The full cohort data

Figure 2 illustrates the fitted models for the full cohort data. Lab tests with significant seasonality are highlighted in red, while those without significance are marked in blue. It indicates that ALP, Monocytes, Neutrophils, Platelets, RBC, and Urea nitrogen/Creatinine had subtle relative seasonality. Among the 11 lab tests showing significant seasonality, Glucose, Neutrophils/100 WBC, and WBC peaked during winter, while others peaked around summer.

**Figure 2.**
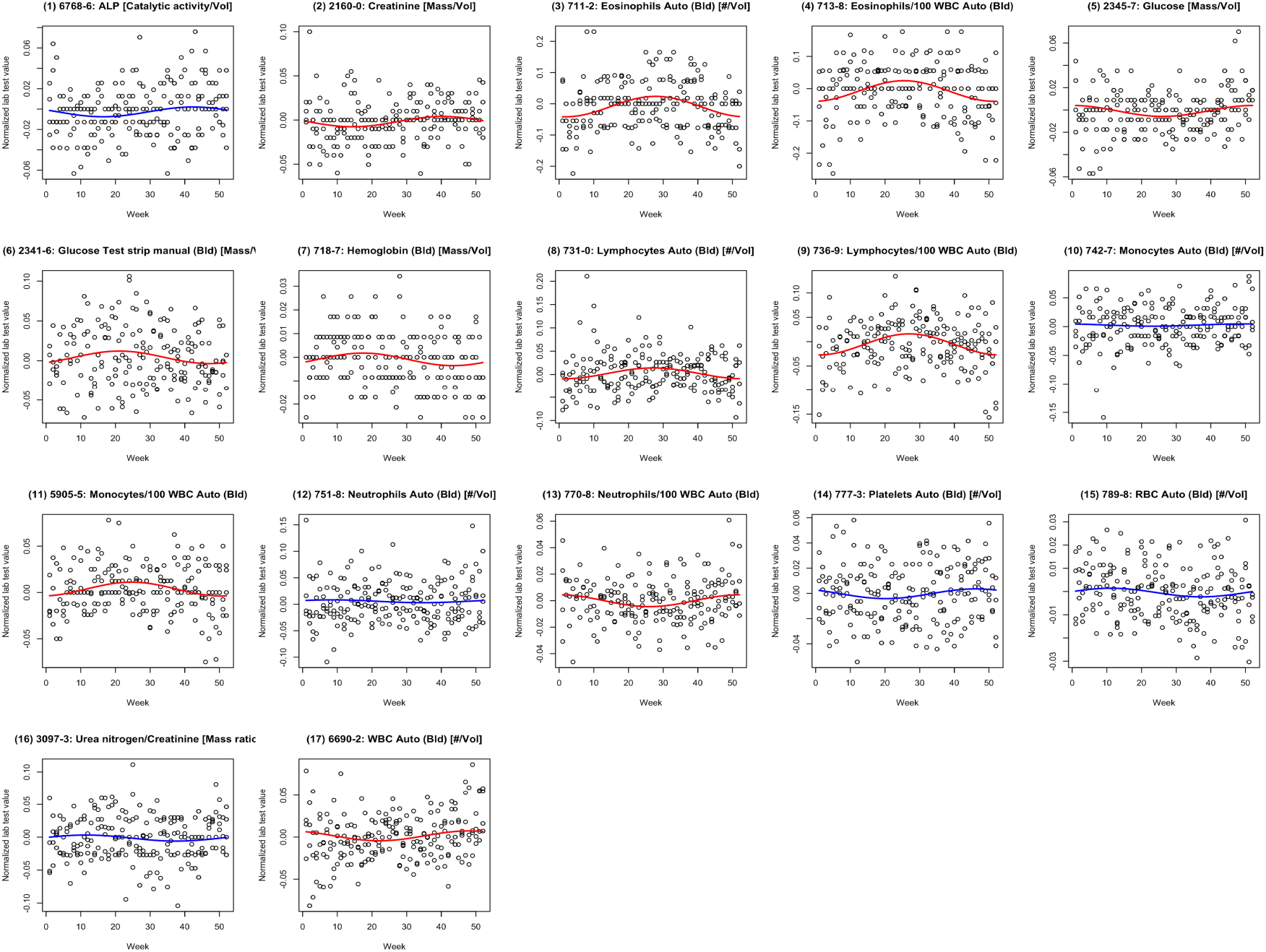
Plots of normalized lab test value versus week for the full cohort. The red lines represent the fitted cosinor models with significant amplitude fits (p<0.05; FDR corrected). The blue lines represent the fitted cosinor model with non-significant amplitude fits (p≥0.05; FDR corrected).

Their significant seasonality findings are summarized in Figure 3(a) using a heatmap, with amplitude and offset fits scaled for clustering. Amplitude shade indicates the magnitude of the fit compared to the largest amplitude fit. For the offset shade, blue represents peak time in summer and red in winter, with intermediate shades indicating spring or fall peaks. Lab tests were grouped into four clusters. Notably, Eosinophils, Eosinophils/100 WBC, and Lymphocytes/100 WBC showed the highest seasonality, with peak values occurring during the summer. Creatinine and Hemoglobin displayed relatively low seasonality, with peak values observed between summer and winter. Glucose test strip manual, Monocytes/100 WBC, and Lymphocytes had moderate seasonality, peaking in summer. Glucose, Neutrophils/100 WBC, and WBC showed low seasonality while peaking during the winter.

**Figure 3.**
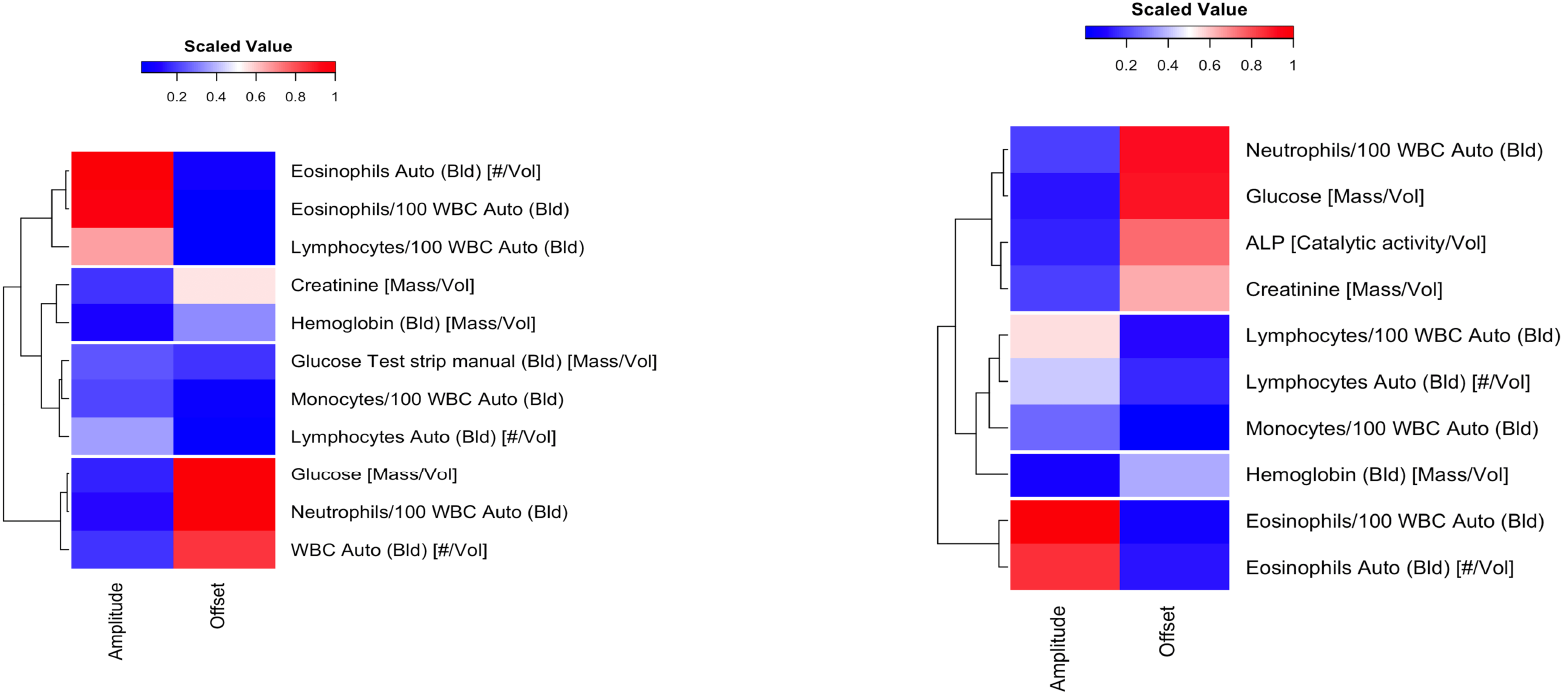
Heatmap for lab tests with significant amplitude fits (p<0.05; FDR corrected) **(a)** for the full cohort data, and **(b)** for the female group data. Amplitude and offset fits are scaled for clustering.

#### (b) Gender stratifications

Acknowledging that males and females may exhibit distinct ranges for certain lab test [24], it is plausible that the seasonality patterns in these two groups could also differ. Thus, we conducted subgroup analysis by fitting models with gender-stratified data. For the male group, only one lab test, WBC, showed significant seasonality, peaking in winter. In contrast, ten lab tests demonstrated significant seasonality for the female group. We utilized a heatmap to summarize their seasonality patterns in Figure 3(b). Specifically, Neutrophils/100 WBC, Glucose, ALP and Creatinine displayed low seasonality, with peak times occurring around winter. Conversely, Lymphocytes/100 WBC, Lymphocytes, and Monocytes/100 WBC had moderate seasonality peaking during summer. Hemoglobin had the least seasonality among all significant lab tests, with peak time in-between. Consistent with the findings from the full cohort data, Eosinophils/100 WBC and Eosinophils displayed the highest seasonality, with peak values occurring during the summer for the female group.

We compared parameter fits of seven lab tests observed in both gender groups. Figure 4(a) presents a comparison of their amplitude fits. Severe seasonal variations were evident in Creatinine, ALP, and Hemoglobin for women, and in WBC and Platelets for men. The difference in their amplitude fits for WBC exceeded 0.007, while RBC showed the most consistent extent of seasonality between the male and female groups.

**Figure 4.**
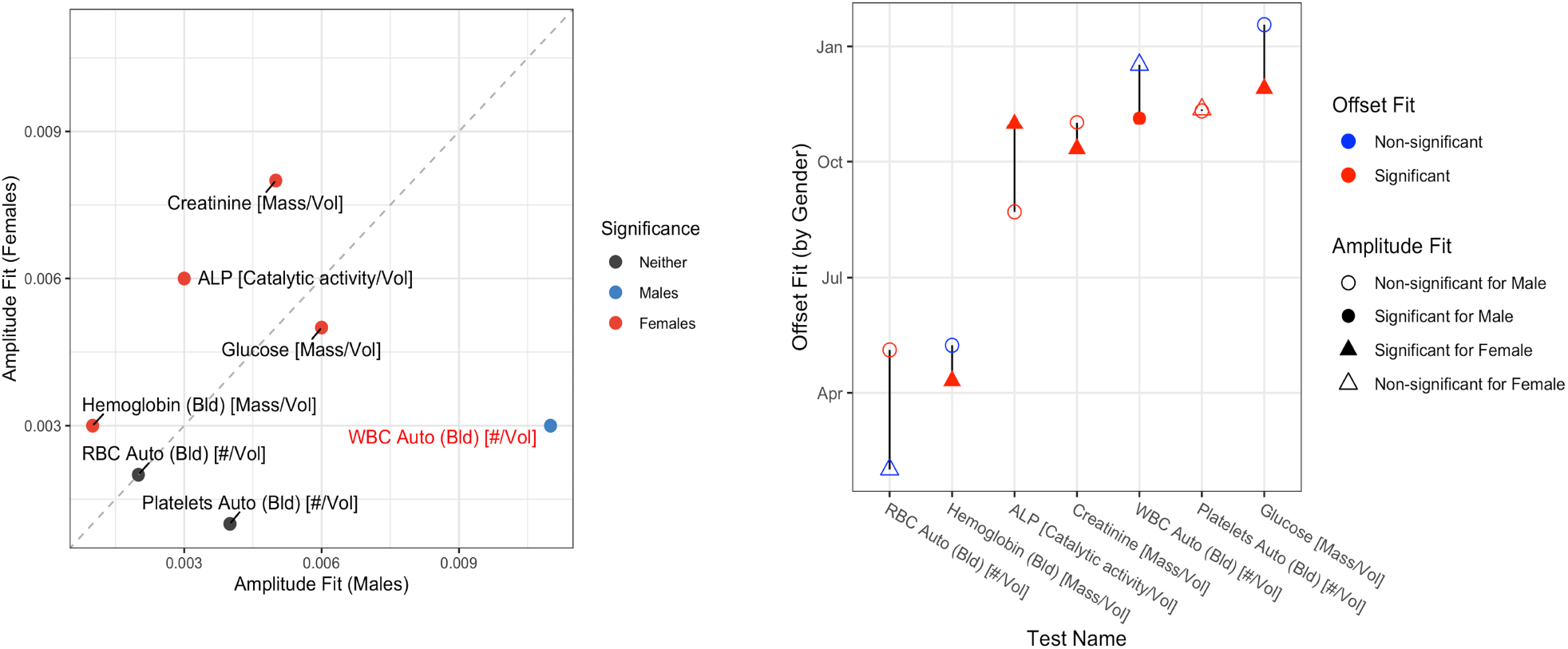
Overview of parameter fits for gender subgroups. **(a)** Amplitude. The lab test is labeled in red if the amplitude fits’ difference between two gender groups exceeds 0.007. The color of each point indicates whether the amplitude fit is significant for either stratum, or neither, accordingly. **(b)** Offset. Point shapes represent genders, with colors indicating offset fit significance, and solidness reflecting amplitude fit significance.

Figure 4(b) compares the time of seasonal peaks for the male and female groups. It shows that lab tests experienced peak values during similar times for both men and women. Interestingly, RBC, the one with the most consistent extent of seasonality in both groups, had the largest difference in peak times. However, since seasonal fluctuations in RBC were negligible for both groups, the potential peak time might vary slightly more due to acceptable randomness in sampling.

#### (c) Age stratifications

Three lab tests exhibited significant seasonal variations for patients under 80 years old, whereas nine showed significant seasonality for patients older than 80. Figure 5 provides an overview of these seasonality patterns for the two age groups. In the group under 80-year-old, all three lab tests had a similar extent of seasonality, with Neutrophils/100 WBC and Creatinine peaking around winter, while Monocytes/100 WBC peaked in summer. For the above 80-year-old group, Platelets, Neutrophils/100 WBC, WBC, and Glucose had the lowest seasonality, with peak times occurring in winter. Lymphocytes, Monocytes/100 WBC, and Hemoglobin also showed low seasonality, but with peaks occurring in summer. Eosinophils, Eosinophils/100 WBC had the highest seasonality, with peaks in summer, consistent with the findings in the full cohort data and the female group.

**Figure 5.**
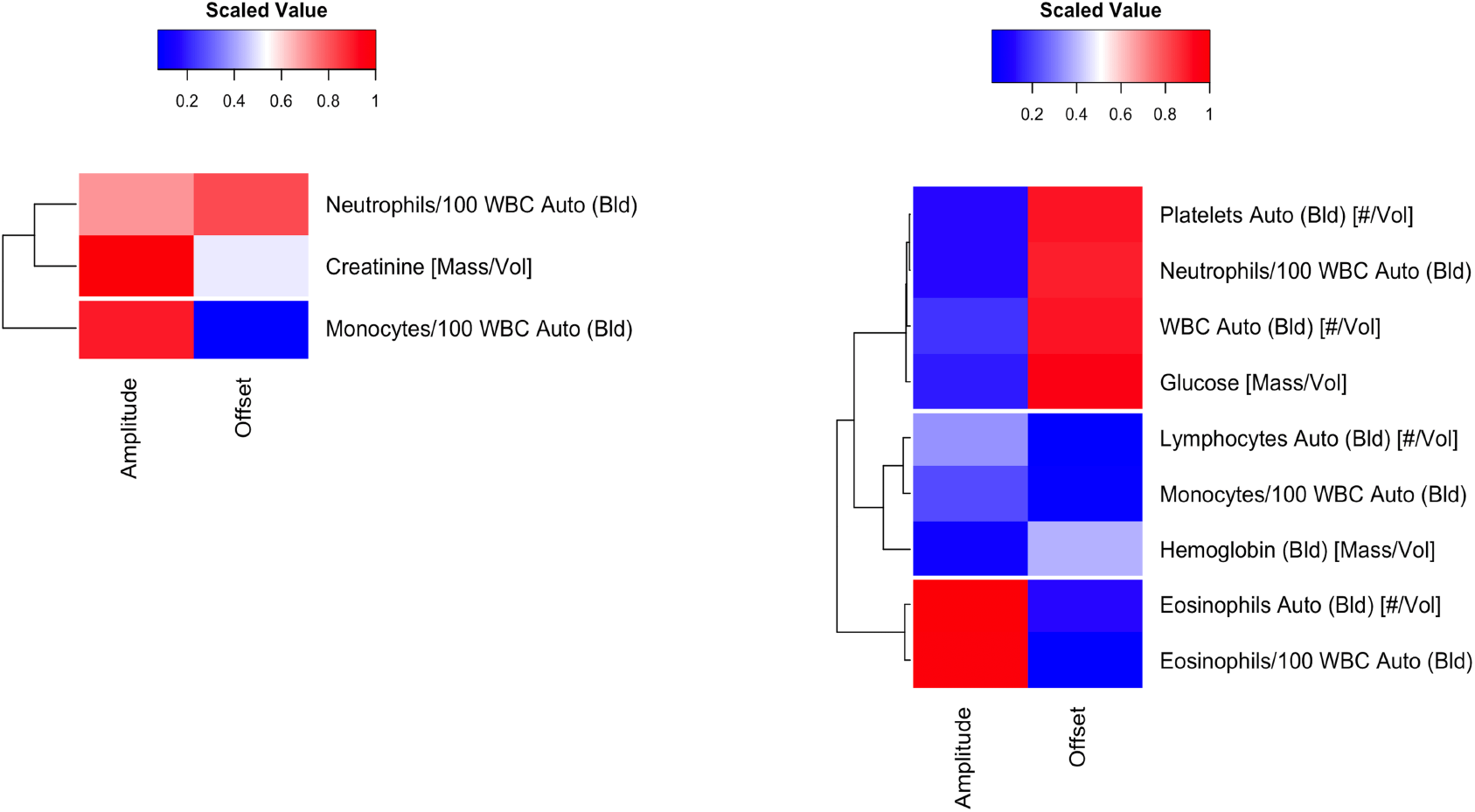
Heatmap for lab tests with significant amplitude fits (p<0.05; FDR corrected) **(a)** for the Age [50-79] group data, and **(b)** for the Age [>=80] group data. Amplitude and offset fits are scaled for clustering.

Figure 6 compares the amplitude fits between two age groups. Lab tests were labeled if the differences in amplitude fits exceeded 0.007. Eosinophils and Eosinophils/100 WBC showed pronounced seasonal variations for the older age group (above 80 years old) at a relative fluctuation of around 0.05, compared to 0.015 for the younger age group (below 80 years old).

**Figure 6.**
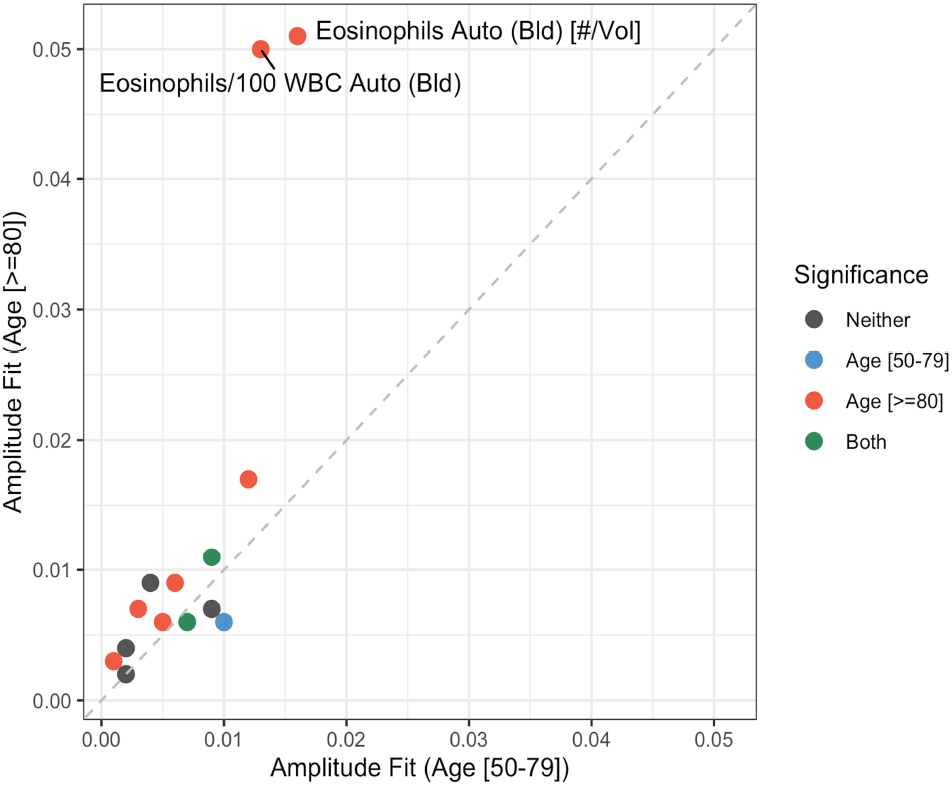
Overview of the amplitude parameter fits for age stratification. Lab tests are labeled if the amplitude fits’ differences between two age groups exceed 0.007. The color of each point indicates whether the amplitude fit is significant for either stratum, or both strata, or neither, accordingly.

Figure 7 compares seasonal peaks for age groups under and above 80. It shows that during summer, patients experienced peak levels for Monocytes/100 WBC, Eosinophils/100 WBC, Lymphocytes, and Eosinophils, regardless of age stratifications. Additionally, for both age groups, Neutrophil/100 WBC, WBC, and Glucose peaked around winter, consistent with the findings observed in the full cohort data and the female group.

**Figure 7.**
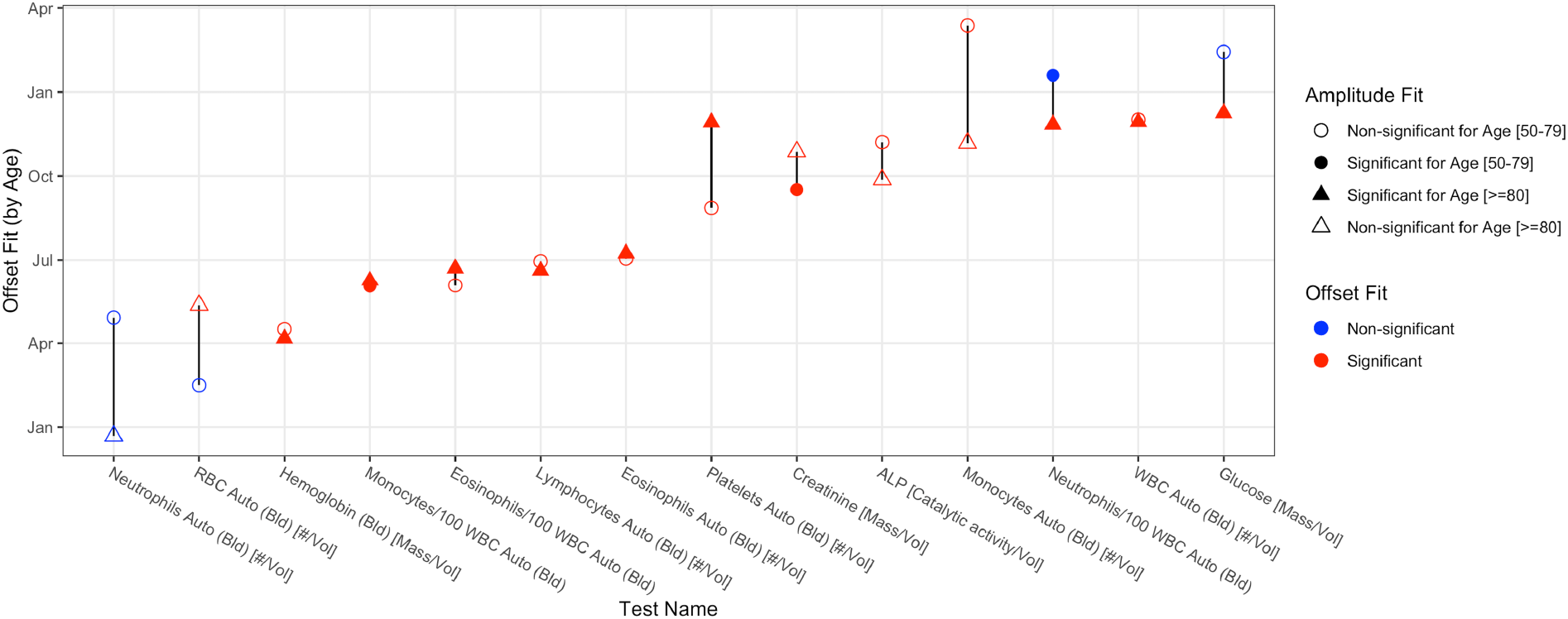
Overview of the offset parameter fits for age stratification. Point shapes represent age groups, with colors indicating offset fit significance, and solidness reflecting amplitude fit significance.

## Discussion and Conclusions

Effective management of Alzheimer’s disease requires a comprehensive approach that includes accurate interpretation of lab tests to monitor and treat co-occurring conditions and complications. This approach supports better disease management, can improve quality of life, and potentially slow the progression of Alzheimer’s disease.

Our study has demonstrated potential seasonal variations in certain lab tests among AD patients in Florida. Glucose, Neutrophil/100 WBC, and WBC levels tend to be higher in winter compared to summer, likely attributed to more indoor activities, shorter daylight hours, and increased carbohydrate intake [25]. The fluctuation of WBC levels has a more pronounced impact on males than on females. Certain leukocyte types, including Eosinophils, Lymphocytes, and Monocytes, typically have elevated levels during summer, possibly due to seasonal respiratory disease and allergens like pollen [11]. The seasonality in Eosinophils appears to be more influential in older AD patients.

Analyzing seasonal patterns in lab test data is essential for healthcare providers and researchers to identify trends, anticipate fluctuations in test values, and tailor patient care accordingly. By recognizing seasonal variations, lab test reference intervals can be adjusted to provide more precise interpretation and diagnosis for individual patients. Additionally, seasonality analysis in lab test data can inform health research strategies, facilitating better data synthesis and enhancing study power. Moreover, many health conditions and exposures have also been documented for their seasonality, including disease incidences, prevalence, and environmental factors [5,26]. Understanding seasonal variations in lab test results would allow researchers to establish connections between risk factors, such as environmental conditions or lifestyle changes, and specific disease incidences through lab tests. By identifying seasonality among risk factors, lab test results, and disease incidence rates, researchers can gain insights into the etiology of diseases. This information is crucial for developing targeted interventions and preventive strategies to mitigate the impact of seasonal risk factors on disease incidence and improve overall public health outcomes.

A few limitations should be noted in this study. Our study analyzed aggregated data, which may result in some information loss. While providing stability, the weekly median of lab test values may potentially counteract seasonal variations compared to individual-level data. Additionally, the number of patients in our study remains insufficient given the breadth of lab tests and the duration of the study period, resulting in many lab tests failing to meet the criteria of at least 100 patients per week and less than 50% missingness. Besides, the cosinor model we utilized assumes the seasonal pattern has a symmetric increase and decrease within one year, which may be too restrictive for seasonal lab tests that do not have a stationary fluctuations [5]. Moreover, it cannot be applied to lab tests with binary or categorical values. In the future, we can explore using the generalized linear model to analyze these types of lab tests.

## Data Availability

The data used in this study cannot be shared due to the privacy concern of the electronic health records.

## Acknowledgments

This project was partially supported by Florida State University Institute for Successful Longevity, the Agency for Healthcare Research and Quality under award number 1R21HS29969, the National Institute on Aging under award number R21AG061431, and the University of Florida – Florida State University Clinical and Translational Science, which is supported in part by the NIH National Center for Advancing Translational Sciences under award number UL1TR001427. VM and SB were supported by the Novo Nordisk Foundation (grants NNF17OC0027594 and NNF14CC0001).

## Notes

### Competing Interest Statement

The authors have declared no competing interest.

### Author Declarations

Florida State University and University of Florida Institutional Review Boards approved this study.

